# Comparison of two measures of cognitive impairment on the 2015 Behavioral Risk Factor Surveillance System

**DOI:** 10.1101/2023.04.06.23288140

**Authors:** Mary Adams

## Abstract

Only selected states have used an optional module to address subjective cognitive decline (SCD) on the Behavioral Risk Factor Surveillance System (BRFSS). This study compares SCD with a core measure of cognitive disability (CDis) that has been included on the BRFSS since 2013. Using 2015 BRFSS data from 35 states that asked the module we report the weighted prevalence of SCD and CDis by demographics, chronic conditions, and risk factors among 150,981 respondents ages 45 years or older. We also compare adults with SCD only, CDis only, both, or neither, plus all those reporting either measure on similar health related measures. In addition, results of logistic regression are presented. Weighted prevalence rates and 95% confidence intervals were 11.5% (11.1%-12.0%) for SCD and 10.6% (10.2%-11.0%) for CDis. Approximately half of those reporting one measure reported the other and about one-third of those reporting either fell into each of the 3 mutually exclusive groups. Comparisons indicated a consistent progression from SCD only, to CDis only, to those with both, for measures of chronic conditions, number of SCD-related risk factors, and poorer health status. Logistic regression results for CDis were more like those for adults with SCD who also reported functional difficulties than for all adults with SCD. Conclusion: Although it does not include a timeframe or capture the exact same respondents as SCD, the core measure of CDis appears to be a useful measure of cognitive impairment and may represent adults more adversely affected than those reporting SCD.

## Introduction

Recognizing that memory problems are often early steps in the development of dementia including Alzheimer’s disease (AD)(1), an optional module was developed for use on the Behavioral Risk Factor Surveillance System (BRFSS) to address cognitive decline (2). Subjective cognitive decline (SCD) was ascertained from the module question “During the past 12 months, have you experienced confusion or memory loss that is happening more often or is getting worse?” The BRFSS collects information from telephone surveys of randomly selected non-institutionalized adults on behaviors related to premature morbidity and mortality (3) and results have been well-validated for many measures (4, 5). Publications using module results (6, 7) have added to the body of knowledge of cognitive decline but are limited to states that chose to ask the module. With many studies suggesting a potential role of primary prevention for AD in the preclinical stage (8-11), surveillance of early cognitive changes becomes vital.

One problem with surveillance of subjective cognitive decline is the lack of standardization in measurement (12, 13). To help meet requirements of the Affordable Care Act (14) for measurement of disability, the Department of Health and Human Services published guidelines (15) that include questions to be used on federal surveys, including the BRFSS. One of those disability questions is “Because of a physical, mental, or emotional condition, do you have serious difficulty concentrating, remembering, or making decisions?” The aim of this study was to compare results for this cognitive disability measure with results for the module measure of SCD to determine the feasibility of using the core measure to study adults with mild to moderate cognitive impairment. BRFSS data from 35 states that asked the SCD module in 2015 would be used.

## Methods

The study used publicly available (16) 2015 BRFSS data from 151,905 (120,726 land line and 31,179 cell phone) surveys of adults ages ≥45 years from the 33 states plus Puerto Rico and the District of Columbia (AL, AZ, AR, CA, CO, DC, FL, GA, HI, IL, IA, LA, MD, MI, MN, MS, NE, NV, NJ, NY, ND, OH, OK, OR, RI, SC, SD, TN, TX, UT, VA, WV, WI, WY, PR) that used the cognitive decline module. Data were weighted to account for the probability of selection, and further adjusted to be representative of the total adult population of each state by age, race, ethnicity, gender, marital status, education, home ownership, and type of phone service. The median survey response rate for these 35 states was 46.6%, ranging from 33.9% to 61.1% (17).

Subjective cognitive decline (SCD) was ascertained from the module question “During the past 12 months, have you experienced confusion or memory loss that is happening more often or is getting worse?” Functional difficulties among those with SCD was determined from responses to two separate questions and included those who responded that, in the past 12 months, confusion or memory loss “always,” “usually,” or “sometimes” (as opposed to “rarely” or “never”) caused them to give up household chores they used to do <u>or</u> interfered with their ability to work, volunteer, or engage in social activities, or both. Respondents that answered “yes” to the following question were deemed to report cognitive disability (CDis):“Because of a physical, mental, or emotional condition, do you have serious difficulty concentrating, remembering, or making decisions?” A four-level measure was also created to compare respondents who reported just SCD, just CDis, both, or neither. There were 2,023 missing values in the 4-level measure (765 due to the SCD measure and 1,258 due to the CDis measure).

Demographic measures included gender, age (45-54, 55-64, 65-74, 75-79, and 80 years and older), self-reported race/ethnicity (non-Hispanic white, Black or African American, Hispanic of any race, American Indian/Alaska Native, and other, which included those of multiple race), education (college graduate, some college, high school graduate, < high school), household income (>$75,000, $50,000-$74,999, $25,000-$49,999, $15,000-24,999, <$15,000, and unknown), reporting a cost barrier to health care (needed to see a doctor in the past year but could not due to cost: yes/no). Health measures included self-reported asthma, arthritis, cardiovascular disease (CVD; heart attack, angina, coronary heart disease, or a stroke), Chronic Obstructive Pulmonary Disease (COPD), disability (any limitation in activity or use special equipment), and health status (fair or poor vs. excellent, very good or good). Other disability measures (e.g. difficulty seeing) have been previously described (18). A composite risk factor index (CDRI)(11) included high blood pressure (ever told), diabetes (ever told except if only when pregnant), obesity (body mass index ≥30 based on self-reported height and weight), sedentary lifestyle (no leisure time physical activity in the past month), current smoking, and depression (ever told they had a depressive disorder). These risk factors were recently shown to be associated with SCD and dementia with dose-response gradients (11). The CDRI was used as a 7 level measure (0-6 risk factors), as an “any” vs. “none” measure, and as a measure distinguishing those with 3 or more risk factors versus <3.

Data analysis was conducted using Stata version 14.1 (StataCorp LP) which accounts for the complex sample design of the BRFSS and used the land line/cell phone weight or the respective version 1 or version 2 weights for the 10 states that used split surveys. Respondents who refused to answer or who answered “don’t know/not sure” were excluded from the analysis involving that measure unless otherwise noted. Prevalence of SCD and CDis and 95% confidence intervals were reported by selected demographic and health-related measures. Respondents in each of the mutually exclusive groups represented by the four-level measure were compared on a similar group of health and demographic measures and selected health outcomes. Also compared were all respondents age 45+ with SCD and all with CDis. Where direct statistical comparisons were not possible, non-overlapping 95% confidence intervals were used to determine statistical significance. Logistic regression was done using either SCD, SCD with functional difficulties, or CDis as the outcome measure and included CVD as a possible confounder (11) because many of the risk factors in the CDRI are also risk factors for CVD.

## Results

For the weighted sample of adults ages ≥45 years, 69.4% were non-Hispanic white, 8.3% were ages 80 and older, 14.6% had less than a high school education, 77.0% reported at least one of the 6 risk factors in the CDRI, 13.9% had CVD, 31.1% were disabled and 23.4% reported fair or poor health. Weighted percentages of adults with SCD and CDis were very similar (Table 1) except that the core measure was associated with gender and SCD was not. A more subtle difference was that the module measure was highest among the oldest age group (≥80 years) while the core measure was similar among all groups except those 65-74 years, where the rate was lowest. Just over half of those reporting each measure also reported the other measure. Among these 35 states, rates of SCD ranged from 6.0% in South Dakota to 16.6% in Nevada while rates for CDis ranged from 6.7% in North Dakota to 16.7% in Mississippi.

**Table 1.** Weighted percentages and 95% confidence intervals (CI) for the core measure of cognitive disability (CDis), and the module measures of subjective cognitive decline (SCD), and functional (Fx) difficulties due to SCD, respondents ages 45 and older, 2015 Behavioral Risk Factor Surveillance System, 35 states, ^a^ N=15,424 for CDis, 16,201 for SCD, and 7,716 for functional difficulties. Sample sizes for SCD and functional difficulties were similar to those listed for CDis.

|  | CDIs |  |  | SCD |  | Fx Difficulties <sup>b</sup> |  |
| --- | --- | --- | --- | --- | --- | --- | --- |
| Group | Percent | 95% CI | Sample Size | Percent | 95% CI | Percent | 95% CI |
| Total | 10.6 | 10.2-11.0 | 150,981 | 11.5 | 11.1-12.0 | 5.8 | 5.5-6.1 |
| **Gender** |  |  |  |  |  |  |  |
| Males | 9.7 | 9.2-10.3 | 60,475 | 11.7 | 11.0-12.3 | 5.6 | 5.2-6.1 |
| Females | 11.3 | 10.8-11.9 | 90,506 | 11.5 | 10.9-12.0 | 5.9 | 5.5-6.4 |
| P value | <.001 |  |  | 0.647 |  | 0.31 |  |
| **Age (years)** |  |  |  |  |  |  |  |
| 45-54 | 11.3 | 10.5-12.1 | 31,911 | 10.8 | 10.0-11.7 | 6.3 | 5.8-7.0 |
| 55-64 | 11.9 | 11.1-12.7 | 44,560 | 11.7 | 11.0-12.4 | 6.6 | 6.1-7.2 |
| 65-74 | 8.0 | 7.4-8.6 | 41,828 | 10.4 | 9.7-11.1 | 4.1 | 3.6-4.5 |
| 75-79 | 10.0 | 8.7-11.5 | 13,538 | 13.4 | 12.0-15.0 | 5.0 | 4.0-6.1 |
| 80+ | 10.8 | 9.8-11.9 | 17,284 | 15.5 | 13.9-17.3 | 5.9 | 5.1-6.8 |
| P value | <.001 |  |  | <.001 |  | <.001 |  |
| **Race/ethnicity** |  |  |  |  |  |  |  |
| White | 9.6 | 9.2-10.0 | 118,707 | 11.3 | 10.8-11.7 | 5.0 | 4.7-5.3 |
| Black | 14.2 | 12.8-15.6 | 13,406 | 13.6 | 12.3-14.9 | 8.5 | 7.6-9.5 |
| Hispanic | 12.1 | 10.8-13.5 | 9,029 | 11.1 | 9.6-12.7 | 7.1 | 6.0-8.2 |
| Am Ind/AN<sup>c</sup> | 24.4 | 19.0-30.7 | 1,675 | 21.1 | 16.4-26.7 | 15.9 | 11.7-21.4 |
| Other | 8.3 | 6.5-10.4 | 5,931 | 10.0 | 7.8-12.7 | 5.0 | 3.7-6.7 |
| P value | <.001 |  |  | <.001 |  | <.001 |  |
| **Education** |  |  |  |  |  |  |  |
| <High School | 20.3 | 18.8-21.8 | 12,382 | 18.1 | 16.6-19.7 | 11.5 | 10.4-12.7 |
| High School | 12.0 | 11.2-12.7 | 43,448 | 12.0 | 11.3-12.8 | 6.4 | 5.9-7.0 |
| Some college | 10.4 | 9.7-11.2 | 40,804 | 11.8 | 11.0-12.6 | 5.7 | 5.1-6.3 |
| College graduate | 4.3 | 3.9-4.7 | 53,888 | 7.4 | 6.9-8.0 | 2.3 | 2.0-2.7 |
| P value | <.001 |  |  | <.001 |  | <.001 |  |
| **Income** |  |  |  |  |  |  |  |
| <\$15K | 27.3 | 25.4-29.2 | 13,711 | 23.0 | 21.1-24.9 | 16.7 | 15.2-18.3 |
| \$15K-\$24,999 | 17.0 | 15.9-18.2 | 21,105 | 16.1 | 15.0-17.3 | 9.9 | 9.0-10.9 |
| \$25K-\$49,999 | 9.7 | 9.0-10.6 | 32,330 | 12.1 | 11.2-13.0 | 5.4 | 4.8-6.0 |
| \$50K-\$74,999 | 6.0 | 5.1-7.1 | 19,830 | 9.5 | 8.3-10.8 | 3.0 | 2.3-3.8 |
| \$75K+ | 4.0 | 3.4-4.6 | 38,511 | 6.2 | 5.6-6.9 | 1.7 | 1.4-2.2 |
| P value | <.001 |  |  | <.001 |  | <.001 |  |
| **Weight status** |  |  |  |  |  |  |  |
| Not overweight | 9.5 | 8.8-10.2 | 44,881 | 11.0 | 10.3-11.8 | 5.4 | 4.9-6.0 |
| Overweight only | 9.5 | 8.9-10.2 | 53,534 | 10.8 | 10.1-11.6 | 5.0 | 4.5-5.5 |
| Obese | 13.2 | 12.5-13.9 | 43,756 | 13.4 | 12.7-14.1 | 7.4 | 6.9-8.0 |
| P value | <.001 |  |  | <.001 |  | <.001 |  |
| **Sedentary lifestyle** |  |  |  |  |  |  |  |
| No | 8.2 | 7.8-8.6 | 105,944 | 9.8 | 9.4-10.3 | 4.4 | 4.1-4.7 |
| Yes | 16.2 | 15.4-17.1 | 44,599 | 15.6 | 14.8-16.5 | 9.2 | 8.6-9.9 |
| P value | <.001 |  |  | <.001 |  | <.001 |  |
| **Ever diagnosed with depression** |  |  |  |  |  |  |  |
| Yes | 36.1 | 34.7-37.6 | 28,542 | 31.2 | 29.8-32.6 | 21.2 | 19.9-22.5 |
| No | 4.9 | 4.6-5.2 | 121,820 | 7.2 | 6.8-7.6 | 2.4 | 2.2-2.6 |
| P value | <.001 |  |  | <.001 |  | <.001 |  |
| **High blood pressure** |  |  |  |  |  |  |  |
| Yes | 13.5 | 12.9-14.1 | 77,190 | 14.1 | 13.6-14.8 | 7.7 | 7.3-8.2 |
| No | 7.9 | 7.4-8.5 | 73,358 | 9.1 | 8.6-9.7 | 4.0 | 3.6-4.4 |
| P value | <.001 |  |  | <.001 |  | <.001 |  |
| **Smoking Status** |  |  |  |  |  |  |  |
| Non-Smoker | 9.0 | 8.6-9.4 | 130,626 | 10.5 | 10.1-11.0 | 4.8 | 4.5-5.1 |
| Current smoker | 20.0 | 18.7-21.3 | 19,449 | 17.8 | 16.5-19.1 | 12.0 | 11.0-13.1 |
| P value | <.001 |  |  | <.001 |  | <.001 |  |
| **Diabetes** |  |  |  |  |  |  |  |
| Yes | 17.0 | 15.9-18.1 | 26,014 | 17.3 | 16.1-18.5 | 10.2 | 9.3-11.1 |
| No | 9.1 | 8.7-9.6 | 124,729 | 10.3 | 9.8-10.7 | 4.8 | 4.5-5.1 |
| P value | <.001 |  |  | <.001 |  | <.001 |  |
| **No. of CDRI risk factors** |  |  |  |  |  |  |  |
| 0 | 2.5 | 2.0-3.1 | 30,995 | 5.0 | 4.3-5.7 | 0.8 | 0.7-1.0 |
| 1 | 5.8 | 5.3-6.5 | 41,908 | 8.2 | 7.5-8.9 | 2.8 | 2.3-3.3 |
| 2 | 11.0 | 10.2-11.9 | 34,467 | 12.4 | 11.4-13.4 | 6.2 | 5.6-6.9 |
| 3 | 18.3 | 17.0-19.7 | 21,177 | 17.3 | 16.1-18.7 | 10.1 | 9.1-11.3 |
| 4 | 30.0 | 28.0-32.1 | 9,454 | 25.8 | 24.0-27.7 | 18.2 | 16.6-20.0 |
| 5 | 43.6 | 39.7-47.5 | 2,520 | 38.7 | 34.9-42.8 | 27.6 | 24.2-31.2 |
| 6 | 61.4 | 50.7-71.1 | 285 | 47.4 | 36.2-58.8 | 43.4 | 32.3-55.2 |
| P value | <.001 |  |  | <.001 |  | <.001 |  |
| **Disability** |  |  |  |  |  |  |  |
| Yes | 26.0 | 25.0-27.1 | 49,581 | 24.1 | 23.1-25.1 | 15.4 | 14.5-16.2 |
| No | 3.6 | 3.3-3.9 | 100,605 | 5.9 | 5.5-6.3 | 1.5 | 1.3-1.6 |
| P value | <.001 |  |  | <.001 |  | <.001 |  |
| **Any CVD** |  |  |  |  |  |  |  |
| Yes | 22.1 | 20.8-23.4 | 22,669 | 23.5 | 22.1-24.9 | 14.1 | 13.0-15.3 |
| No | 8.5 | 8.1-8.9 | 126,679 | 9.5 | 9.1-10.0 | 4.4 | 4.1-4.7 |
| P value | <.001 |  |  | <.001 |  | <.001 |  |
| **Health Status** |  |  |  |  |  |  |  |
| Fair or poor | 27.3 | 26.1-28.6 | 33,400 | 25.5 | 24.3-26.7 | 16.8 | 15.8-17.8 |
| Good or better | 5.4 | 5.1-5.8 | 117,108 | 7.2 | 6.8-7.6 | 2.4 | 2.2-2.7 |
| P value | <.001 |  |  | <.001 |  | <.001 |  |
| **Serious difficulty seeing** |  |  |  |  |  |  |  |
| Yes | 40.3 | 37.5-43.1 | 9,326 | 33.9 | 31.3-36.6 | 26.2 | 23.7-28.9 |
| No | 8.5 | 8.2-8.9 | 139,395 | 9.9 | 9.5-10.3 | 4.3 | 4.1-4.6 |
| P value | <.001 |  |  | <.001 |  | <.001 |  |
| **Report the other measure (SCD or CDis)** |  |  |  |  |  |  |  |
| Yes | 52.4 | 50.5-54.4 | 15,985 | 57.5 | 55.6-59.4 |  |  |
| No | 5.0 | 4.8-5.3 | 131,878 | 6.1 | 5.8-6.4 |  |  |
| P value | <.001 |  |  | <.001 |  |  |  |
| **State (includes DC & PR)** |  |  |  |  |  |  |  |
| AL | 15.6 | 14.4-17.0 | 5,116 | 12.9 | 11.8-14.2 | 7.8 | 6.9-8.9 |
| AZ | 11.0 | 9.8-12.3 | 5,179 | 13.5 | 12.2-14.9 | 6.6 | 5.7-7.7 |
| AR | 14.7 | 12.9-16.7 | 3,725 | 16.2 | 14.3-18.3 | 9.2 | 7.7-11.0 |
| CA | 8.8 | 7.2-10.7 | 1,946 | 11.7 | 9.8-14.0 | 4.8 | 3.7-6.2 |
| CO | 8.9 | 7.8-10.2 | 4,079 | 10.8 | 9.6-12.2 | 4.6 | 3.7-5.6 |
| DC | 10.2 | 8.1-12.8 | 2,722 | 12.5 | 10.1-15.4 | 6.2 | 4.3-8.8 |
| FL | 10.8 | 9.3-12.4 | 3,110 | 11.3 | 10.0-12.9 | 6.3 | 5.2-7.6 |
| GA | 12.1 | 10.6-13.7 | 2,956 | 14.1 | 12.5-15.9 | 7.6 | 6.4-9.1 |
| HI | 7.4 | 6.4-8.5 | 4,458 | 9.0 | 7.9-10.2 | 4.4 | 3.6-5.3 |
| IL | 7.5 | 6.4-8.7 | 3,369 | 9.7 | 8.5-11.0 | 4.8 | 3.9-5.9 |
| IA | 8.5 | 7.5-9.6 | 4,193 | 9.3 | 8.2-10.4 | 3.2 | 2.7-3.9 |
| LA | 16.2 | 14.5-18.0 | 2,953 | 14.7 | 13.1-16.5 | 8.2 | 6.9-9.6 |
| MD | 9.5 | 7.9-11.5 | 4,422 | 10.8 | 9.1-12.8 | 4.9 | 3.9-6.2 |
| MI | 12.7 | 10.8-14.9 | 1,930 | 12.1 | 10.3-14.0 | 5.5 | 4.3-6.9 |
| MN | 8.0 | 7.3-8.6 | 10,725 | 8.7 | 8.1-9.4 | 3.3 | 2.9-3.8 |
| MS | 16.7 | 15.2-18.4 | 4,343 | 12.9 | 11.5-14.5 | 8.5 | 7.3-9.9 |
| NE | 8.4 | 7.5-9.5 | 5,788 | 9.5 | 8.5-10.7 | 4.3 | 3.6-5.1 |
| NV | 11.1 | 8.8-13.9 | 1,865 | 16.6 | 13.7-19.9 | 7.4 | 5.5-10.0 |
| NJ | 8.3 | 7.4-9.4 | 7,434 | 9.2 | 8.2-10.3 | 4.5 | 3.8-5.3 |
| NY | 10.2 | 8.8-11.7 | 3,817 | 11.2 | 9.8-12.7 | 5.6 | 4.5-6.9 |
| ND | 6.7 | 5.7-8.0 | 3,398 | 10.0 | 8.7-11.4 | 4.6 | 3.7-5.7 |
| OH | 10.4 | 9.3-11.5 | 8,398 | 10.8 | 9.7-11.9 | 5.5 | 4.8-6.3 |
| OK | 14.7 | 12.8-16.9 | 2,390 | 13.7 | 11.8-15.8 | 7.9 | 6.4-9.6 |
| OR | 13.2 | 11.7-14.9 | 3,291 | 13.2 | 11.7-14.8 | 6.6 | 5.4-8.1 |
| RI | 10.3 | 9.0-11.8 | 4,149 | 11.6 | 10.2-13.2 | 6.2 | 5.1-7.6 |
| SC | 13.3 | 12.3-14.5 | 7,748 | 12.2 | 11.2-13.2 | 7.2 | 6.4-8.1 |
| SD | 7.7 | 6.5-9.2 | 5,067 | 6.0 | 4.9-7.2 | 2.7 | 2.1-3.5 |
| TN | 14.1 | 12.6-15.9 | 3,961 | 13.3 | 11.9-15.0 | 7.7 | 6.5-9.2 |
| TX | 10.9 | 9.4-12.6 | 4,438 | 13.1 | 11.4-15.0 | 7.4 | 6.1-9.0 |
| UT | 8.8 | 7.5-10.4 | 3,035 | 11.1 | 9.5-12.9 | 4.0 | 3.1-5.1 |
| VA | 8.3 | 7.4-9.3 | 5,638 | 8.9 | 8.0-9.9 | 4.3 | 3.7-5.1 |
| WV | 14.7 | 13.4-16.0 | 3,995 | 9.9 | 8.9-11.1 | 5.3 | 4.6-6.2 |
| WI | 8.6 | 7.4-9.9 | 3,852 | 10.9 | 9.6-12.4 | 4.4 | 3.5-5.4 |
| WY | 8.9 | 7.6-10.4 | 3,911 | 11.2 | 9.8-12.8 | 4.7 | 3.7-5.8 |
| PR | 13.4 | 12.1-14.9 | 3,580 | 6.6 | 5.7-7.7 | 5.2 | 4.3-6.2 |
|  | <.001 |  |  | <.001 |  | <.001 |  |

Abbreviations: CDis: cognitive disability; CI: confidence interval; CVD: cardiovascular disease; Fx: Functional; SCD: subjective cognitive decline.
<sup>a</sup> States are: AL, AZ, AR, CA, CO, DC, FL, GA, HI, IL, IA, LA, MD, MI, MN, MS, NE, NV, NJ, NY, ND, OH, OK, OR, RI, SC, SD, TN, TX, UT, VA, WV, WI, WY, PR.
<sup>b</sup> Functional difficulties; determined from responses to 2 questions and include those who responded that, in the past 12 months, memory problems always, usually, or sometimes (as opposed to rarely or never) caused them to give up household chores they used to do or interfered with their ability to work, volunteer, or engage in social activities, or both.
<sup>c</sup> American Indian or Alaska Native

Table 2 compares the approximately equal number of respondents reporting just SCD, those with only CDis, and those reporting both, and omits the majority of respondents reporting neither measure who consistently reported the most favorable results. These results highlight the differences between only SCD, only CDis, and the combination, showing that in general, there was a progression in that order in terms of adverse effects. This was despite the fact that adults with just SCD tended to be older than those in the other groups. Results for all adults ≥45 years with either SCD or CDis (i.e. non-mutually exclusive categories) were consistent with both Table 1 and the 4-way results. Although only the 57.5% of respondents with CDis who also reported SCD were asked about functional difficulties, 76.7% of all respondents reporting functional difficulties due to SCD were respondents reporting CDis (not shown). Adults with CDis reported a higher mean number of the 6 risk factors for cognitive decline (11) than those with SCD (2.6 vs. 2.3) and were also more likely to report any of the risk factors or 3 or more of them. The exception to the general progression from SCD to CDis was noted for most of the chronic conditions, where rates were similar for all respondents reporting SCD and all those reporting CDis, where, as noted, there was about a 50% overlap.

**Table 2.**
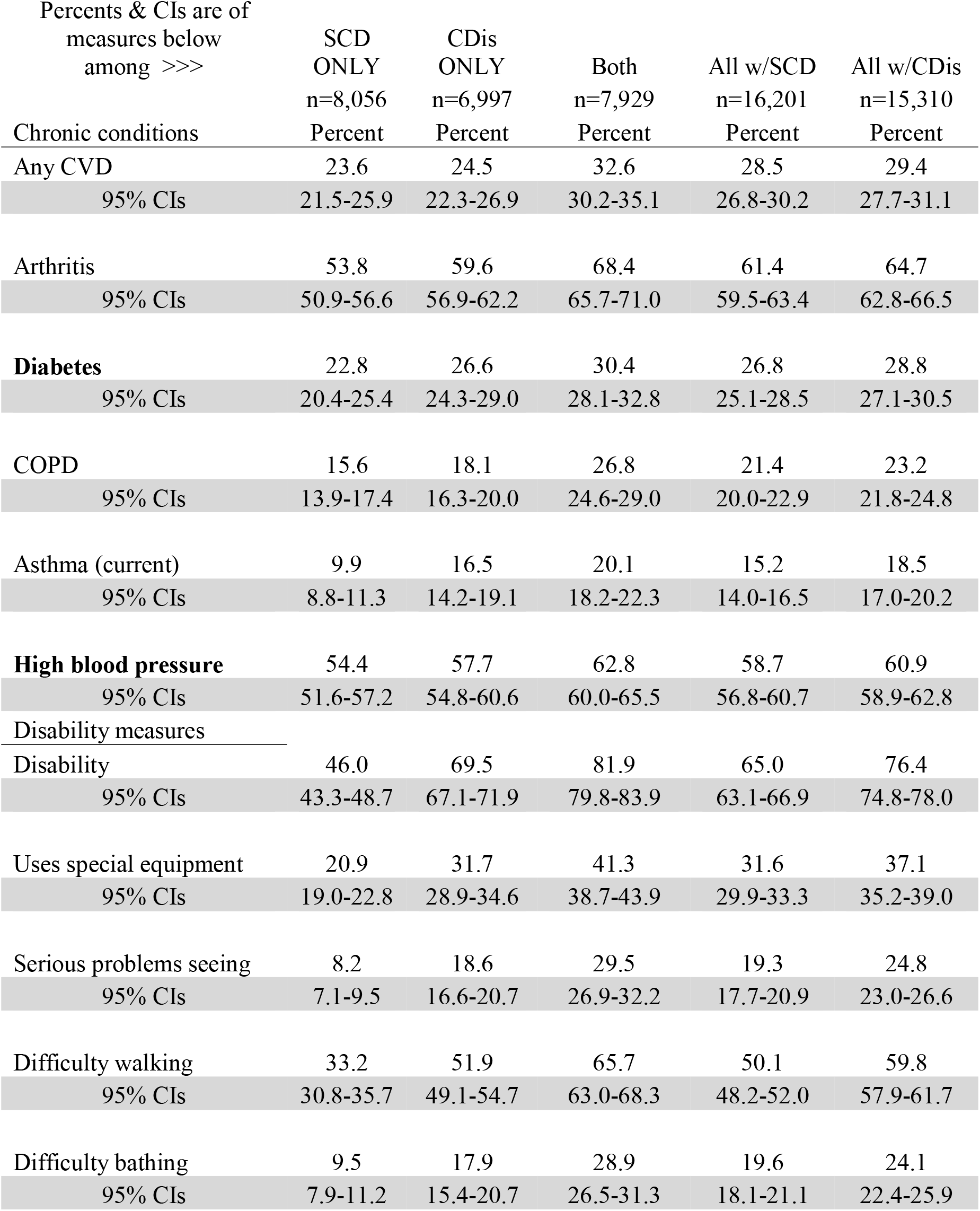
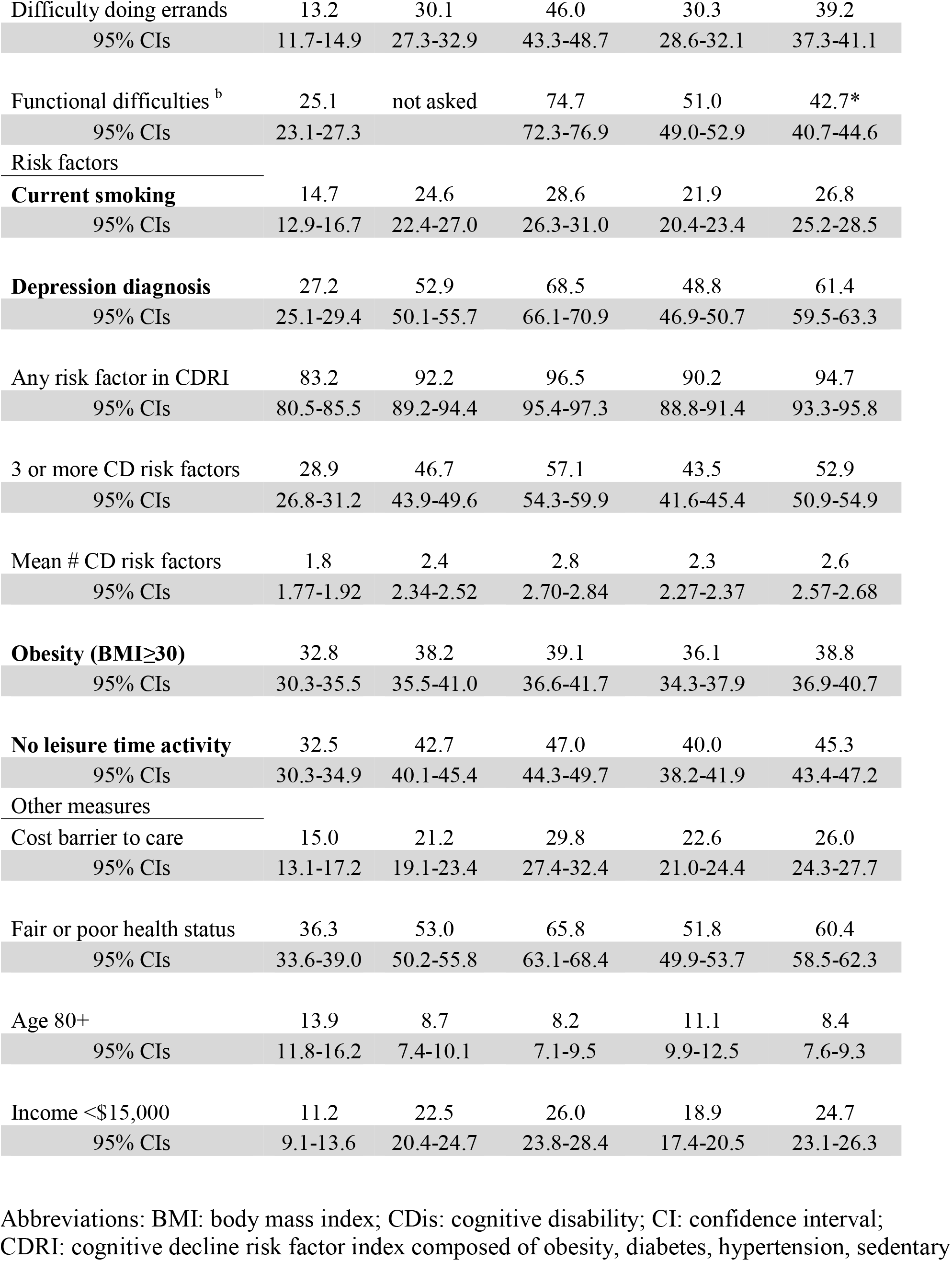
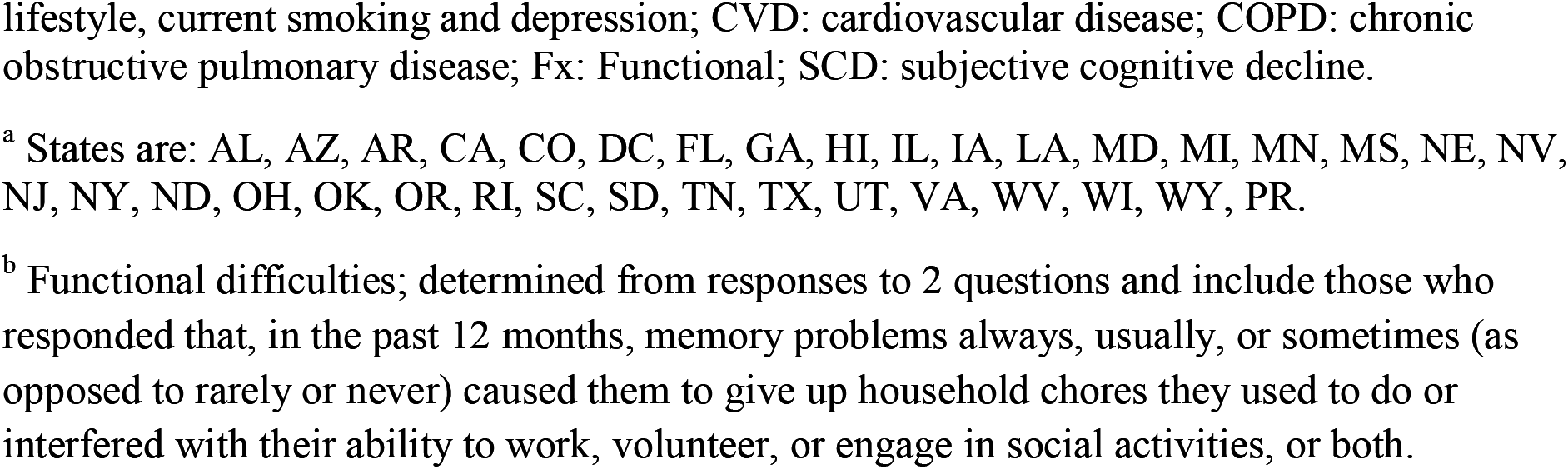
Comparison of 3-level measure with mutually exclusive categories of just subjective cognitive decline (SCD), just serious cognitive disability (CDis), or both, plus columns for all respondents with SCD and all with CDis, 2015 Behavioral Risk Factor Surveillance System, 35 states ^a^, respondents ages 45 years and older. Measures in bold are the six risk factors in the cognitive decline risk index (CDRI). Results for those with neither measure have been omitted.

Results of logistic regression (Table 3) confirmed the unadjusted gender difference for CDis but also showed attenuated associations for age, income, and education compared with the unadjusted results shown in Table 1, especially for SCD. In each model, the highest adjusted odds ratio (OR) was for adults reporting all 6 risk factors, with ORs of 7.6 for SCD, 24.5 for CDis, and 24.8 for functional difficulties associated with SCD, and clear dose-response gradients shown. With the exception of gender differences, results for CDis and SCD with functional difficulties were more similar to each other than either one was to SCD. Without the addition of “difficulty seeing” to the model (not shown), ORs for American Indians/Alaska Natives vs. non-Hispanic whites were >1.45 for cognitive disability and SCD with functional difficulties but was not significant for just SCD.

**Table 3.**
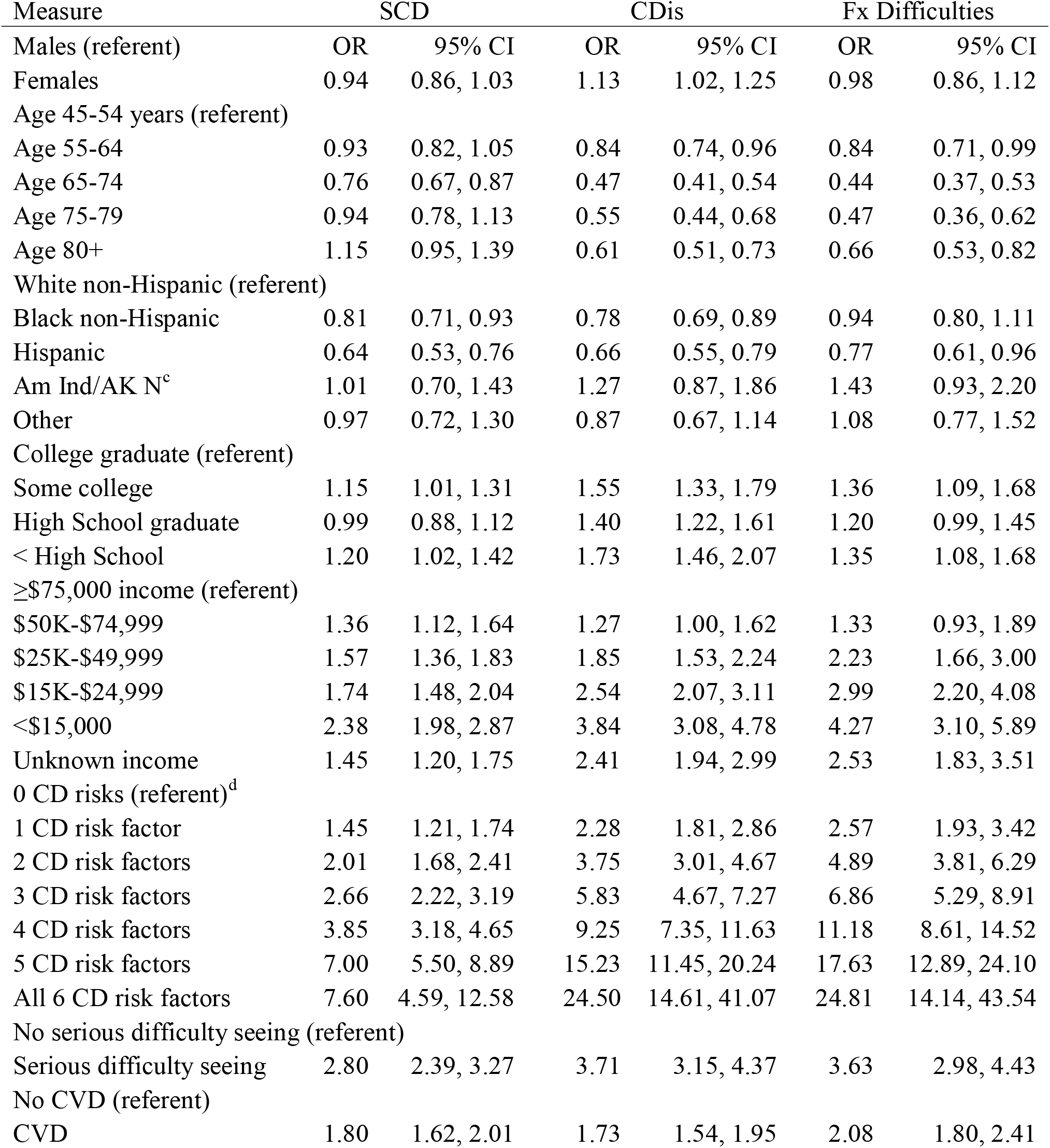
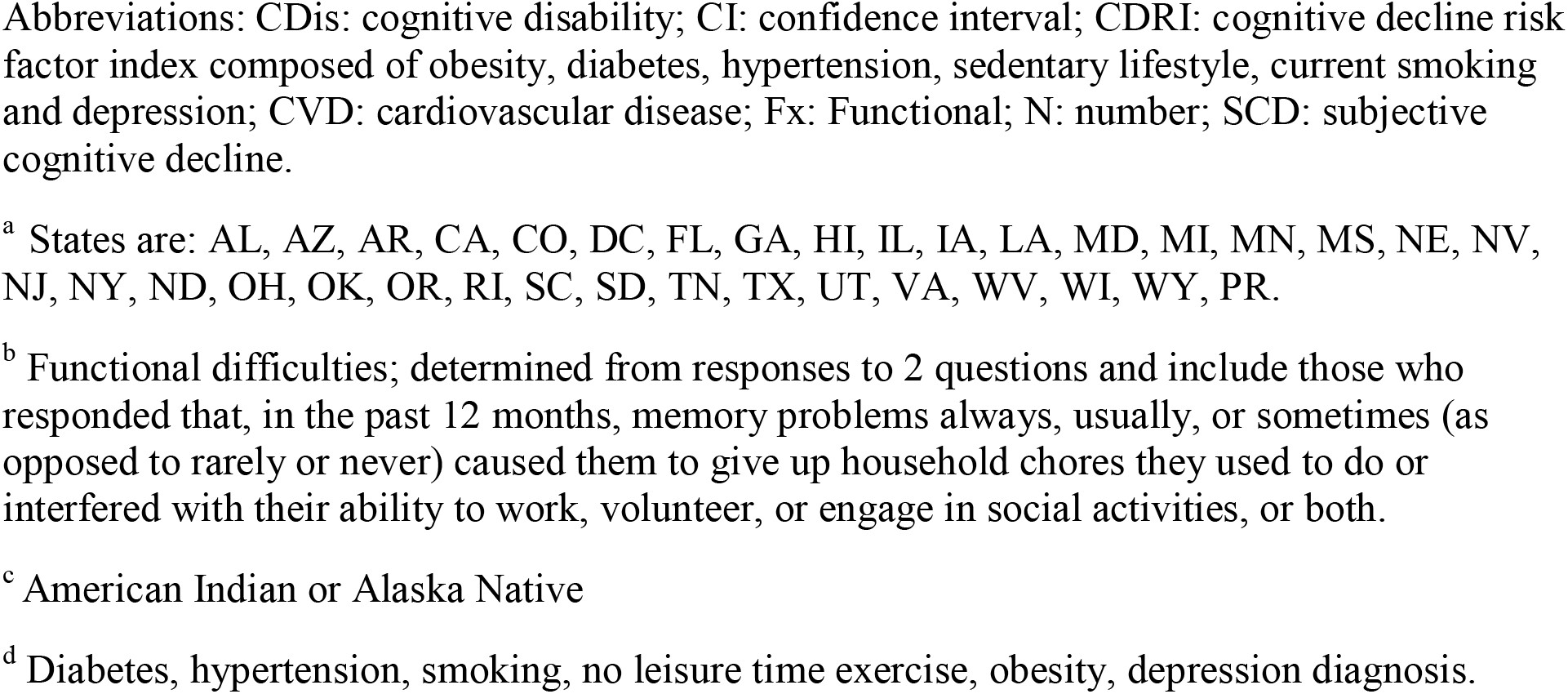
Adjusted odds ratios (ORs) and 95% confidence intervals (CI) for selected outcomes, adults ages ≥ 45 years, adjusted for all measures listed; 2015 Behavioral Risk Factor Surveillance System, 35 states.^a^ Total number (N) for each model ≥ 135,000; Cognitive decline (SCD) N= 16,201; Cognitive disability (CDis) N=15,310; Functional (Fx) difficulties^b^ N= 7,716.

## Discussion

Prevalence estimates for the cognitive disability measure and the module measure of SCD are similar (10.6% and 11.5% respectively) although they do not capture exactly the same respondents. The measures overlap by about 50% and the disability measure, but not the module measure, is more prevalent among women. From results of the mutually-exclusive 4-level measure, it appears that respondents reporting only SCD are least affected by memory problems or confusion, followed by those with only CDis, with those reporting both being the worst off. Those results are consistent with results indirectly comparing all respondents with SCD with all those reporting CDis, although these latter results are less dramatic and not always statistically significant.

Currently there is no standard measure of subjective cognitive decline, but the importance of a pre-clinical measure of Alzheimer’s disease has been recognized. The Subjective Cognitive Decline Initiative (SCD-I) Working Group was established to help define measures to be used in studies of pre-mild cognitive impairment (12). The key items considered for a measure of SCD were 1) as a subjective measure, no confirmation by cognitive testing was required; 2) “cognitive” could refer to any area of cognition and was not limited to memory; and 3) “decline” refers to a worsening of cognition. As part of their work, 34 measures used in 19 studies (13) were examined in terms of context, language, cognitive domain, timeframe, and other factors.

Neither of the measures used in this current study appears to have been included. Among the 34 measures only 15 included a timeframe for change while several addressed degree of severity. The Working Group did not rule out measures without a timeframe but recommended that studies document whether or not the measure included the concept of decline. They also suggested that measures which do not include a timeframe should be considered measures of “cognitive impairment” rather than “decline”. Thus it appears that while the core disability measure (CDis) does not measure “decline”, it is appropriate for use as a measure of “cognitive impairment.” The phrase “serious difficulty” in the question may indicate a higher degree of severity compared with cognitive decline (12), which is consistent with the findings in this study. Further evidence for the CDis measure being appropriate as a measure of cognitive impairment is indicated by the dose-response gradients shown between all three study outcome measures and the CDRI. Both unadjusted and logistic regression results indicate that the associations between SCD, SCD with functional difficulties, and the CDis measure with the 6 risk factors in this study are similar to those shown for SCD and functional difficulties in an earlier study (11). The results for the module measures confirm those earlier results while similar results for CDis add support for its use as a measure of cognitive impairment.

The CDis measure offers potential advantages compared with the module measure of SCD. Not only is it asked on the core of the BRFSS so is available for all states, but other federal surveys such as the Health Interview Survey (19) and American Community Survey (20) include the same question. These additional surveys expand the potential for studying early cognitive changes. Also, starting in 2015 the module measure was restricted to ages 45 and older while respondents of all ages are asked the core disability measure. While follow-up questions asked on the cognitive decline module are not asked of respondents reporting cognitive disability (unless they also report SCD from the module), they could always be asked as state added questions. Because not all SCD eventually leads to AD (12), the CDis measure can also be used to study other possible causes of cognitive impairment especially in younger respondents.

Adjusted results from logistic regression deserve special mention. Unadjusted results for all three measures are highest among American Indians/Alaska Natives but this result is not confirmed in the logistic regression results presented here. However, as noted in Results, if the measure of “difficulty seeing” is excluded from the model, odds ratios for this racial group are ≥1.45 for CDis and SCD with functional difficulties. While more study might help to understand factors that could explain this finding, the fact remains that unadjusted rates for all three measures are significantly higher among American Indians/Alaska/Natives. Thus it appears appropriate to consider this racial group as being at higher risk for cognitive impairment compared with non-Hispanic whites for these 35 states.

Because the BRFSS random selection process excludes entire households if the selected adult is reported to be unable to respond to a telephone survey, some selected adults with dementia or cognitive impairment might result in their household being excluded. Earlier versions of the cognitive decline module included questions for other (non-respondent) adults in the household with SCD. These other adults may or may not have been able to respond to the survey, as that criterion was not applied to them. Results from 2011 module data for 21 states (21) that included proxy responses for non-respondent adults with SCD found that 39% of all household adults with SCD were non-respondents. Compared with respondents with SCD, these non-respondents were much more likely to report functional difficulties, getting informal care and treatment for their SCD and to have been diagnosed with dementia. Thus it is important to note that these 2015 study results only represent respondents capable of answering a telephone survey.

There are several limitations to this study. Persons in households with no telephones are excluded although those with only cellular telephones are now included. Data are self-reported and reliability and validity can vary for measures tested (4, 5). The validity of the cognitive decline module measures is not known, nor has testing been done among respondents with cognitive impairment. However, cited BRFSS validity testing would have included respondents with cognitive decline who were deemed capable of responding to the survey. We purposely avoided adding measures to the logistic regression model that might show reverse causality (e.g. disability) but expect that adding other variables could affect results as was noted for American Indians/Alaska Natives. Because many of the cognitive decline risk factors are implicated in other chronic diseases, similar dose-response results could be expected for a variety of outcomes. However, conditions that do not share these risk factors, such as skin cancer, do not show these results (11). Due to the variability of results across these 35 states (Table 1), the generalizability of the results is unknown.

In conclusion, it appears that the measure of cognitive disability on federal surveys is an appropriate measure of cognitive impairment, bearing in mind that it can’t measure “decline” because it lacks a timeframe. Adults reporting CDis may be somewhat more affected by their impairment compared with respondents reporting SCD from the BRFSS cognitive decline module. Future studies using the data for all 50 states and available territories could include confirmation of results for the 6 risk factors and CDis in all states to determine generalizability. Further examination of racial differences in the measure for all 50 states might confirm whether American Indians/Alaska Natives are at increased risk for cognitive impairment. Demographic differences in cognitive impairment might lead to better targeting of interventions. Other potential risk factors for SCD could be studied. States that have never asked the cognitive decline module could reap immediate benefits by using the CDis measure to estimate cognitive impairment in their state.

## Data Availability

All data produced in the study are either available in the paper or on request.

http://www.cdc.gov/brfss/annual_data/annual_2015.html

## Acknowledgments

No financial support was received for this work.

